# Client and Health Care Provider Perspectives on Point-of-Care Testing for Sexually Transmitted Infections in Community Pharmacies in Uganda

**DOI:** 10.64898/2026.04.02.26350027

**Authors:** Annet A. Onzia, Adelline Twimukye, Johan H. Melendez, Matthew M. Hamill, Peter Kyambadde, Agnes Kiragga, Yukari C. Manabe, Rosalind Parkes-Ratanshi

## Abstract

**Introduction:** The World Health Organization (WHO) recommends testing for sexually transmitted infections (STIs), but laboratory-based and rapid or point-of-care (POC) testing are often unavailable and unaffordable, especially in low-resource settings, leading to empiric (usually antibiotic) treatment. Community pharmacies (CPs) are often the first point of contact for persons with symptoms of STIs, where pharmacists dispense treatment without diagnostic testing or prescriptions. This study evaluated clients, providers, and policymakers’ perspectives on POC testing for STIs in CPs for targeted treatment.

**Methods:** We nested a qualitative study into a study of participants seeking both STI and non-STI treatments in CPs. They were tested for HIV, syphilis, trichomonas, chlamydia, and gonorrhea using both rapid POC tests and a central reference lab. A purposive sample of 50 participants from September 2020 to June 2022 consented to participate in in-depth and key informant interviews. Data were analyzed thematically using an inductive approach.

**Results:** Clients (n=35), health care providers (n=9), and policy makers (n=6) highlighted the benefits of POC tests for HIV and STIs at CPs, including affordability, accessibility, and ensuring convenience. The impact of POC testing for STI diagnosis and treatment was promoting behavioral change, rapid results turnaround time, leading to faster treatment access compared to conventional laboratory methods, and supporting sustainable antimicrobial resistance (AMR) control. Barriers to POC testing included a lack of awareness among clients and health workers, inadequate privacy and space, long wait times, unclear self-sample collection instructions, stigma around HIV testing, and reluctance to test for STIs beyond HIV. To address these, participant recommendations included raising STI awareness, providing more explanation of test results, increasing test access, addressing stigma, provider training, and ensuring a sustainable supply chain for testing kits.

**Conclusions:** POC testing for STIs and HIV in CP settings was found to be highly acceptable to both pharmacy clients and providers. Integrating POC testing in CPs could be beneficial for national STI management programs. If the existing barriers are addressed, POC tests could improve accessibility to STI diagnostics and facilitate better linkage to care.

## BACKGROUND

Sub-Saharan Africa (SSA) bears a significant burden of sexually transmitted infections (STIs), which are increasing globally, and remain under-researched (1, 2). STIs may cause serious adverse health outcomes, including pelvic inflammatory disease, infertility, congenital infections, and neonatal death. Non-HIV STIs increase the risk of HIV transmission and continue to fuel the ongoing HIV epidemic in the region (3).

The syndromic approach to STI management misses asymptomatic cases, leads to continued transmission, and may result in overtreatment, contributing to antimicrobial resistance (4). To address the rising global STI prevalence, there is an urgent need to shift to an etiologic approach (4). Point-of-care tests (POCTs) offer greater flexibility compared to conventional laboratory methods and can be used in rural and community settings, providing quick results that aid faster decision-making and improve care efficiency (5, 6). Lateral flow assays, in particular, are widely accepted in low- and middle-income countries (LMICs) (7–9); they are the backbone of HIV and syphilis diagnostic testing in these settings.

In Uganda, poor access to STI services in public healthcare has led many clients to seek care in private and informal sectors, such as community pharmacies (CPs) and traditional healers (10). CPs, often called retail pharmacies, are dedicated stores or drug outlets where licensed pharmacists sell prescribed and compounded medications. They may offer a convenient and discreet setting for STI testing and treatment, with more flexible hours that often include round-the-clock availability. In addition, many provide broader public health services, including antiretroviral treatment and sexual and reproductive health (SRH) care (11). As trusted health care providers, pharmacy staff offer private consultations in settings already involved in sexual health, including condom sales and emergency contraception (11, 12). Whilst there are about 3595 licensed pharmacies in Uganda, which comply with Ministry of Health guidelines (13), there are many unlicensed premises where prescription-only antibiotics for STI treatment are often available over the counter, leading to an increased risk of incorrect or inadequate treatment, which contributes to antimicrobial resistance (14–16). Whilst some CPs have been approved by the Ministry of Health to provide antiretroviral therapy, many operate in isolation with limited integration into the national health care system (17, 18). Whereas data from high-income countries show the feasibility and acceptability of STI/HIV testing in CPs (19), there are limited data on its implementation in LMICs like Uganda.

Understanding the experiences and attitudes of CP users, providers, and policy makers towards sexual health service provision in CPs is crucial for evaluating the potential for consistent, high-quality services (20), exploring the potential for integration into the national health care system, and identifying training needs (21). We undertook a nested qualitative study within a prospective cohort study of persons seeking medication and other services at CPs in Kampala and Wakiso districts, Uganda [23]. We explored clients’, providers’, and policymakers’ perspectives on the awareness, acceptability, and satisfaction with POC STI/HIV testing in CPs.

## METHODS

### Study Design

We conducted a nested qualitative study with a purposive sample of 50 participants, employing in-depth and key informant interviews, enrolled in a prospective cohort study. The cohort study was conducted from September 2020 to June 2022 (22); 450 participants seeking both STI and non-STI treatment in CPs provided blood samples for HIV/syphilis testing using the SD Bioline (Abbott, Illinois, US). Self-collected vaginal swabs for *Trichomonas vaginalis* (TV) were tested using the OSOM^®^ Trichomonas Rapid Test (Sekisui, San Diego, USA) POCT. *Neisseria gonorrhoeae* (NG) and *Chlamydia trachomatis* (CT) testing of self-collected vaginal swabs in women and urine in men, with GeneXpert (Cepheid, HBDC, Maurens-Scopont, France). All parent study participants received sexual health education, including counselling about STI/HIV prevention and treatment. Partner notification for those with positive tests for STIs included in-person or through phone calls or through an interactive voice response tool (Call for Life) (23). All participants were followed up and re-tested for all STIs, except HIV, on days 30 and 90 to ascertain treatment response, symptom resolution following the baseline visit (when applicable), and partner notification for persons who tested positive for any STI at previous visits. In this manuscript, POC testing refers to the HIV, syphilis, and TV testing that were performed and resulted within the premises of the CP, while NG and CT tests are considered near-POC tests, as their samples were transferred to the main laboratory and results received between 12 and 24 hours later.

### Study settings

The study was conducted in 18 private CPs located in Kampala city and the neighboring peri-urban district of Wakiso in Uganda, with a population of approximately 3.8 and 3.5 million people, respectively(24). By March 2020, there were approximately 353 licensed pharmacies in the five divisions of Kampala and about 66 in Wakiso District. (24).

### Sampling size estimation

Using maximum variation sampling, we purposively selected 54 participants for in-depth interviews (IDIs) and key informant interviews (KIIs) across five divisions of Kampala and municipalities in Wakiso District, Uganda. Of these, four declined to participate due to time constraints. We conducted 35 IDIs with CP clients enrolled in the parent study. These included persons seeking emergency contraception (n=3), STI treatment (n=18), both emergency contraception and STI treatment (n=1), and those seeking care for other conditions not classified in the aforementioned categories (n=13). Both male and female participants were enrolled, and recruitment within each category continued until data saturation was achieved (**Table 1**).

**Table 1.** Participant characteristics for the in-depth interviews.

| Variable | Frequency (%), n=35 |
| --- | --- |
| Sex |  |
| Female | 20 (57.1) |
| Male | 15 (42.9) |
| Age (Median (IQR)) | 29 (25-40) |
| Highest level of education |  |
| None | 1 (2.9) |
| Primary | 14 (40.0) |
| Secondary | 15 (42.9) |
| Tertiary | 3 (8.6) |
| University | 2 (5.7) |
| Marital status |  |
| Married | 18 (51.4) |
| Separated | 6 (17.1) |
| Single | 11 (31.4) |
| Reason for Visiting the Pharmacy |  |
| Asymptomatic – other reason | 13 (37.1) |
| Emergency Contraception | 3 (8.6) |
| Symptomatic of STI | 19 (54.3) |
| Occupation |  |
| Accountant | 1 (2.9) |
| Boda rider | 4 (11.4) |
| Business | 12 (34.3) |
| Car Park attendant | 1 (2.9) |
| Casual worker | 1 (2.9) |
| Washes clothes | 1 (2.9) |
| House maid | 2 (5.7) |
| House wife | 2 (5.7) |
| Mason | 1 (2.9) |
| Mason & boda boda rider | 1 (2.9) |
| Mobile money agent | 1 (2.9) |
| Plumber | 1 (2.9) |
| Student | 2 (5.7) |
| Support staff | 1 (2.9) |
| Tailor | 1 (2.9) |
| Taxi driver | 1 (2.9) |
| Teacher | 1 (2.9) |
| Long-distance truck driver | 1 (2.9) |

*One(n=1) participant visited the pharmacy for both STI symptoms and emergency contraception and was considered under the STI symptoms category*

We also conducted 15 KIIs with health care providers and stakeholders, including pharmacists, dispensers, representatives from the Ministry of Health National AIDS Control Program, the Pharmaceutical Society of Uganda, and the National Drug Authority. Key informants were selected for their expertise in STI management and/or pharmacy operations. Interviews took place either at the participating CPs or at the Infectious Diseases Institute clinic to ensure privacy (**Table 2**).

**Table 2.**
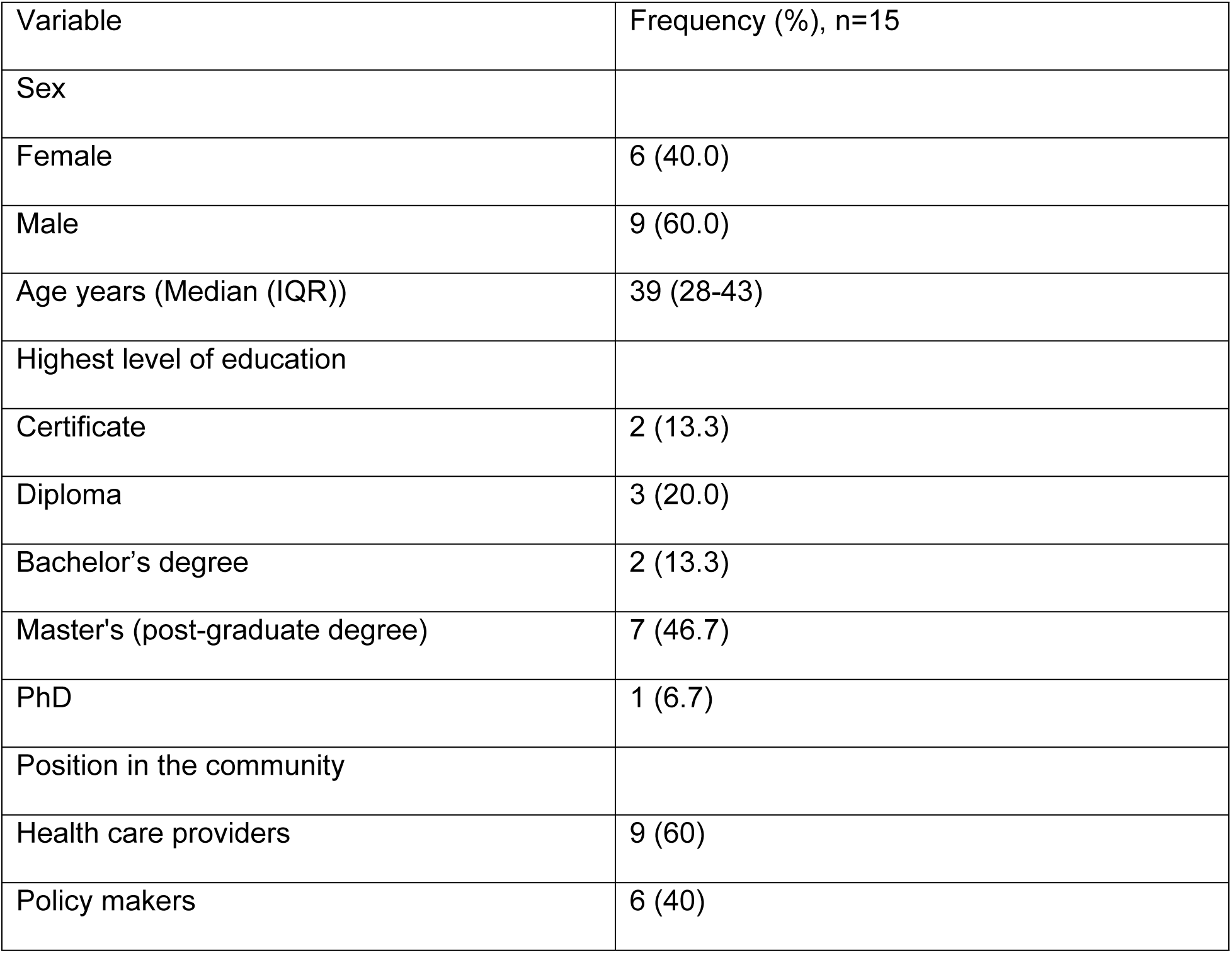
Characteristics of participants for the key informants’ interviews.

| Variable | Frequency (%), n=15 |
| --- | --- |
| Sex |  |
| Female | 6 (40.0) |
| Male | 9 (60.0) |
| Age years (Median (IQR)) | 39 (28-43) |
| Highest level of education |  |
| Certificate | 2 (13.3) |
| Diploma | 3 (20.0) |
| Bachelor's degree | 2 (13.3) |
| Master's (post-graduate degree) | 7 (46.7) |
| PhD | 1 (6.7) |
| Position in the community |  |
| Health care providers | 9 (60) |
| Policy makers | 6 (40) |

### Ethical considerations

The study was approved by the Institutional Review Board (IRB) at The Joint Clinical Research Center (JC1820) in Uganda and the Johns Hopkins University (IRB00279241). The Uganda National Council of Science and Technology (HS1274ES) provided overall regulatory and administrative oversight and approval regarding the conduct of study activities. We obtained further permission from the district health offices of the two districts in Kampala and Wakiso, as well as from the proprietors of the individual pharmacies where the study activities took place. Written informed consent was obtained from all participants before enrollment. For participants deemed illiterate or semi-illiterate, we used a thumbprint in the presence of an impartial witness. Participation in the study was entirely voluntary, and whoever wished to withdraw was allowed to do so without any repercussions. All pharmacy staff signed a confidentiality agreement with the study to ensure the confidentiality and privacy of study participants enrolled from their facilities. All study participants received time compensation of ∼$10 USD, as guided by the IRB. All participants’ personal information and identifiers were removed and anonymized during analysis and reporting using alphanumeric codes to ensure confidentiality. All data were password-protected and stored on encrypted computers and locked cabinets.

### Data Collection

Data were collected by study team members experienced in qualitative data collection in both English and Luganda (the predominant local language). Both IDIs and KIIs used semi-structured guides in participants’ preferred language, lasting 30 to 60 minutes. Interviews were audio-recorded and supplemented with written notes. The topic guides, based on Sekhon et al.’s acceptability framework (25) and the Ugandan Ministry of Health’s STI Management Guidelines (26), covered areas such as knowledge, perceived benefits and risks, barriers, and attitudes towards POC STI/HIV testing in CPs. Data collection was staggered, allowing emerging results from IDIs to be verified during subsequent KIIs and continuing until saturation was reached.

### Data Analysis

The audio data were translated into English, if recorded in another language, transcribed, coded, and thematically analyzed to address the study objectives. To ensure transferability, we used a thick description of the findings (27). Data accuracy and completeness were verified by an independent review of all transcripts. Analysis was conducted using NVivo version 14 software and a thematic approach. A flexible coding framework was developed to facilitate inter-district comparisons and accommodate emerging themes. Each transcript was read multiple times, and relevant text segments were coded. Similar codes were grouped to form themes, which helped develop narratives. Contradictory data were treated as potential different viewpoints and analyzed further to validate them. Validated data enriched insights on the research topic. To ensure rigor, each transcript was independently coded, compared, and discussed by four study team members. Emerging themes were reviewed and agreed upon by the study team. In this manuscript, we report key phrases and verbatim quotes from study participants’ experiences and perspectives according to the Consolidated Criteria for Reporting Qualitative Research (COREQ) (28).

## RESULTS

Among 35 participants in the IDIs, the median age was 29 (IQR 25-40) years, with the majority female, 57.1% (n=20), married 51.4% (n=18); 14.3% (n=5) had post-secondary education (**Table 1**). The reasons for visiting the pharmacy were as follows; 54.3% (n=19) sought care for STI symptoms, 8.6% (n=3) wanted emergency contraception, and 37.1% (n=13) had visited for other reasons besides the two aforementioned. For the KIIs, the median age was 39 (IQR 28-43) years, 60% (n=9) were male, and the majority, 66.5% (n=10), had post-diploma education (**Table 2**).

The analysis identified four main themes related to POC testing for STIs/HIV at CPs. The first theme revealed participants’ general perceptions about the benefits of POC testing at CPs, including the quick turnaround time, quality of services, and counseling services offered. The second theme assessed the impact of POC testing for STIs/HIV at CPs. The third emerging theme identified the social and structural barriers to POC STI/HIV testing at CPs, while the fourth theme identified recommendations by participants for the implementation of POC STI/HIV testing at CPs. These themes cut across various levels at the individual, interpersonal, health systems, and policy levels.

### Theme 1: Benefits of POC testing at CPs

#### Good quality and comprehensive services offered

Our study participants highlighted the good quality and comprehensive nature of the services, which included counselling, education on HIV/STI prevention and treatment, medication provision, and follow-up calls. The health education and counselling, which focused on prevention, testing, and treatment of STIs/HIV, emphasized the importance of knowing one’s health status and making informed decisions, as emphasized by one of the participants who tested positive for HIV.

> *“… I felt scared a bit, but then since I received good counselling, I said let me enroll for treatment so that I can practice positive living…But when she [nurse] calls me…I get encouraged and I conclude that, when I am on treatment, I can get well”*-- IDI, Male, symptomatic.

#### Reduced costs of POC testing

Participants appreciated the availability of free POC testing provided by the parent study, which contrasted with the high costs associated with conventional STI testing. This made testing more accessible, especially for those who previously lacked resources or were deterred by the high costs of consultations and laboratory fees.

> *“Their services were good, and yet they were free. Exorbitant costs are usually associated with traditional STI testing…. consultation before you get in…then lab fees…”* --IDI, Female, asymptomatic.

#### Ease of accessibility

CPs were viewed as convenient, being well-positioned and accessible in both urban and rural settings. They were seen as trusted spaces for testing, with clients appreciating their proximity and the regulatory standards that ensured quality.

> *“…community pharmacies are very accessible…there is some kind of trust between the community and the pharmacy. They are quite well positioned… to penetrate deeper areas. Go to any upcoming town, and you will find a pharmacy there.”* --KII, MOH technical advisor for vaccines and therapeutics.

#### Ease of use of POC tests

POC tests were considered convenient and easy to use, with some health care workers feeling empowered to perform tests for pharmacy clients within the pharmacy, which contributed to faster diagnoses and treatment management.

> *“…we can just do it [test] ourselves, find out the truth, then we manage”*-KII, Pharmacy dispenser.

### Theme 2: Impact of the POC testing for STIs at the CPs

#### Sexual health education

Participants reported that the education they received with POC testing encouraged changes in sexual behavior. Testing encouraged clients to make healthier lifestyle choices, such as committing to one sexual partner and adopting safer practices.

> *“…My biggest lesson has been that it is always important to test and know your HIV status because it will impact your overall sexual relations…after this testing, I chose to stick to my officially introduced wife and I openly told her, I am done with [other] women, let me settle down with you and we raise our children”* -- IDI, Male, asymptomatic.

> *“I am not supposed to have many sexual partners. You need to have one partner, and encourage him to be checked like you.”*- IDI, Female, symptomatic.

#### Etiological diagnosis for targeted management

Participants noted that POC testing led to accurate diagnoses and targeted treatment, which could result in recovery from curable STIs and improved health outcomes, compared to the standard of care syndromic management for STIs available at other facilities. Participants acknowledged the significance of rapid testing before treatment to ensure the appropriate medications were prescribed.

> *“In the context of antimicrobial resistance, this could go miles in reducing the time someone accesses effective treatment, so I see them to be the future.”* --KII, Male, MOH Consultant

#### Quick results turnaround time

The quick turnaround time of POC tests was seen as a major advantage, particularly for those with limited time who might not visit public health facilities. The turnaround time of results made it easier for clients to begin treatment promptly. This also led to encouragement for their sexual partners to get tested if found positive for STIs/HIV, implying that they wanted to avoid reinfection by encouraging their partners to get tested and treated for STIs/HIV following POC testing.

> *“It saves a lot of time. Someone will be out in 15 minutes…they will tell you that I have no time to go to the lab. I am too busy. But if they come to you and you tell them, just give me 10 minutes. I carry out the test, they are [so much] willing.”* --KII, Pharmacy dispenser.

> *“I took it upon myself to inform my partner to get tested after me…the tests are easy and quick.”* --IDI, Male, Symptomatic

### Theme 3: Barriers to STI testing in CP

#### Lack of awareness about POC tests among clients and providers

Both clients and health workers often lacked awareness of POC testing for STIs apart from HIV, since the usual practice was to describe symptoms to pharmacists and receive medication without being tested. The majority of participants had attended the CP to buy medicine to treat various non-STI symptoms. Some health workers recognized the need to educate both the public and practitioners about the accuracy and benefits of POC testing, which could increase its uptake if available.

> *“No, I had never heard about it. I just bumped into it.”* --IDI, Male, Symptomatic

> *“There is a knowledge gap…if the public could know that point of care is accurate…quick, then they would take it up…that should start with medical workers”* -- KII, male, pharmacist.

#### Sample collection challenges

Many participants noted that CPs lacked privacy, as tests were conducted and results provided in open spaces, discouraging honest communication and increasing fear. There was insufficient space and/or a designated bathroom to conduct tests and engage clients comfortably, which meant that participants used public or other shop toilets.

> *“…the pharmacy was in the open and people kept passing…I was a bit scared …it did not have privacy.”* -- IDI, Female, symptomatic.

> *“Yes, because you have to go into people’s toilets…You might find that it is locked…at the trading center, you had to first pay to use the toilet.”*--IDI, Female, Symptomatic

Some male clients found the instructions for self-sample collection unclear, leading to discomfort and possible inaccuracies in sample collection. The process of collecting samples was difficult for some participants, despite clear instructions given, which deterred many from testing.

> *“At some point, I didn’t know how deep I was to push the swab…She [nurse] didn’t tell me how far I can put it”* --IDI, Female, symptomatic.

#### Inadequate training of pharmacy providers

Pharmacists expressed concern around lack of training around POC testing, raising concerns about the quality of the testing process and the validity of results.

> *“Do we have the expertise to collect the right sample? In the hospital, you have a lab tech…I don’t think, as a pharmacist, we have sufficient training, our training tells us so much…how to collect a sample…simple things like a prick, an HIV test that is okay”* --KII, Female, pharmacist.

#### Long queues and waiting times at CPs

Our participants observed that CPs are busy, and therefore, there are often long queues that discourage testing and waiting for results. In some cases, requiring additional laboratory confirmatory testing (especially for HIV), this caused anxiety and deterred people from testing.

> *“The worry about standing in long queues for long in the open deters people… although I had negative results for all the STIs…when is she [nurse] calling me, why is she not?”* --IDI, Female, symptomatic.

#### Stigma associated with HIV testing

The stigma associated with HIV testing, particularly the fear of receiving positive results, was a significant barrier to POC testing. Some participants were afraid of the emotional and social consequences of knowing their HIV status, such as isolation or self-harm in an environment with no counsellors.

> *“Some people are afraid to know their status. Some others might take poison if they find out that they have HIV.”* --KII, Male, pharmacy technician.

#### Concerns about the diagnostic performance of POC tests

Health workers raised concerns about the accuracy and reliability of POC tests, particularly regarding false positives and negatives

> *“The challenge with POCs is sensitivity and specificity… the false negatives and false positives.”* --KII, Male, Consultant medical officer

#### Potential risks of Antimicrobial resistance

Participants expressed concern that placing POC test kits in pharmacies could lead to misuse, as pharmacists may lack the clinical training needed to use them properly, potentially contributing to antimicrobial resistance.

> *“Why not put these kits and support the clinics which are designated to do real patient care…other than equipping the drug outlets which are run by a little bit lower cadre … the placement of POC test kits in pharmacies is likely to perpetuate antimicrobial resistance (AMR) as it encourages pharmacists to undertake clinical work for which they have inadequate training.”* --KII, MOH STI control program.

The profit-driven nature of pharmacies was seen as a potential conflict of interest, where financial incentives might take precedence over patient care, leading to inappropriate tests and/or prescriptions.

> *“It happens a lot in pharmacies… dispensers are under pressure, overlooking health to focus on money”*-IDI, Female, asymptomatic.

### Theme 4: Recommendations for implementing POC Testing at CPs

#### Raising awareness of POC testing and reducing stigma

Participants recommended that more sensitization about the availability of POC testing for HIV and STIs is needed using contextually relevant methods. This sensitization needs to be matched with counselling to reduce stigma around STI/HIV testing, addressing fears, shame, and misinformation during community sensitization campaigns.

> *“It would be good if things like flyers are put there…Messages that say testing is for free on this and that day…. the use of loudspeakers going through communities encouraging people to get information about that sexually transmitted disease, signs and symptoms.”* --IDI, Female, symptomatic.

Participants recommended that additional public health sensitization around antimicrobial resistance and misuse of antibiotics in both humans and animals is urgently required.

> *“We may need to create messages that are easy to understand, for the local population, so they know that it is not good to go and buy antibiotics without a prescription for themselves or their animals… Drugs have been misused not only in human beings but in animals. So, the emergence of resistance is the biggest challenge around, misuse of drugs, under doses.”* --KII, medical officer.

#### Contextually appropriate POC testing

To increase access to POC testing, participants recommended that testing points be accessible and convenient within local communities, for example, with dedicated testing days to reduce queues and waiting times.

> *“I think they should get some days in a week and say these are the days they test.”* –IDI, Female, Asymptomatic

#### Address privacy concerns at CPs

To enhance privacy at CPs, participants recommended creating safe spaces within or outside CPs to facilitate open, confidential communication between clients and health practitioners, further encouraging POC testing.

> *“We get like a tent, that is placed on the side, so that the client and the health worker…you are just in your conversation.”*—IDI, Female, Symptomatic

> *“I think point of care is a cheap option compared to the conventional laboratory one…It also saves a lot of space, as I have said, I could create some small room there that basically runs point of care.”*—KII, Male, Pharmacist

#### Use of alternative non-invasive samples for testing

Participants further recommended that easy-to-collect samples, like urine, saliva, or small blood drops, be used to increase uptake of POC testing at CPs.

> *“…it has to be samples that are culturally, technologically friendly, like saliva, urine.”* --KII, Male, STI Control Unit

Further, participants recommended making the POC tests easier to use by translating instructions into local languages to enable self-sample collection and results interpretation, as asserted by one of the participants.

> *“…let them even try to bring it in Luganda [local language] to make it easy for someone to read it”* --KII, Female, comprehensive nurse/dispenser.

#### CP POC testing to support Antimicrobial stewardship

The use of POCTs to support antimicrobial stewardship was recognized and welcomed by providers and policymakers. Participants suggested that the government strengthen the enforcement of regulations to prevent the over-the-counter sale of STI treatments by non-registered pharmacies. Supervision could help enforce laws to prohibit the sale of antibiotics for STIs without proper testing and prescription, as this contributes significantly to AMR. Others emphasized the need for pharmacists to refer clients to qualified clinicians for appropriate treatment and for engagement with traditional healers.

> *“No one goes to check at my pharmacy to see if the prescriptions dispensed and antibiotics brought in…no regulation is the major stumbling block.”* –KII, Male, Pharmacist

> *“Even the herbalists have contributed to AMR. They mix their herbs…get some doxy, get some streptomycin, and boil…So when someone says, “I can treat kabotongo[syphilis],” they are more than sure; they know what they put inside there…in that herbal, they have laced it with conventional medicine. Of course, it can’t be stable because they have boiled it, mixed it with other things…if we can involve the traditional medicines…like opinion leaders, traditional council…we can let them know about the dangers of mixing the herbals with the conventional medicine.”* --KII, Male, National Drug Authority

POC tests were recognized as supporting a move from syndromic management of STIs to testing-based treatment with more control of AMR.

> *“If the ministry found personnel to visit the community pharmacies to ensure drugs are dispensed on prescription…following up the prescription books…regularly, it will force dispensers…to tell clients to test before prescription.”* -- KII, Male, Consultant medical officer.

#### Regular review and evidence-based update to local guidelines

Updating policies to reflect current evidence and best practices to effectively address emerging challenges, especially around AMR, is important. Regular training on new policies was emphasized.

> *“The guidelines should be informed by actual local AMR surveillance and not necessarily just changed because of literature adopted anywhere.”* –KII, Male, STI Control Unit

> *“We should really have this constant training to update our knowledge”*-KII, Female, pharmacist.

#### Ensuring the constant availability of supplies

Participants recommended that POC testing kits and treatment need to be consistently available in CPs for a sustainable and trustworthy service.

> *“…the supply chain has to be guaranteed and sustainable”* --KII, Pharmacist, vaccines, drugs supply chain.

However, not all our participants welcomed CP POC testing. Some challenged the concept of CP providing POC testing, as they felt this should be done only in clinics.

> *“…these pharmacies, their mandate…are not licensed to treat…so why not equip the nearby clinics and then they get drugs from the pharmacy…Why not put these kits and support the clinics…which are designated to do real patient care…other than equipping the drug outlets which are run by a little bit lower cadre?”* --KII, Medical officer consultant.

## DISCUSSION

This study is among the first to document the high acceptability and satisfaction with POC testing for HIV and STIs among providers, policy makers, and individuals seeking treatment for STIs and other health conditions at community pharmacies in Africa. This study highlights a key role for CPs in increasing access to STI services. Participants viewed CPs as valuable alternatives to traditional syndromic care in health facilities, and cited pharmacists’ professional training as valuable to conduct POC testing. These findings are similar to those reported in South Africa, where community pharmacies were recognized for their potential to alleviate the burden of public health facilities in providing routine HIV testing and treatment, especially to underserved populations (29).

Before the study, both clients and providers had limited awareness of POC or near-POC testing, mainly due to inadequate community sensitization. These findings align with previous studies in various SSA settings, which report low awareness of HIV/STI POC testing leading to low uptake of services (30). We also demonstrate high acceptability and satisfaction with POC testing among CP users and health care providers. Participants highlighted the ease of sample collection and rapid result interpretation as key facilitators of testing. Same-day treatment was highly valued, as it encouraged initial testing uptake, treatment completion, and partner notification. These results are consistent with previous studies that document high acceptability of POC STI testing because it saves time and cost (7, 31). However, some health care providers expressed dissatisfaction with test instructions being available only in English, which limited client comprehension, findings consistent with a study in India where participants felt disempowered to use POC diagnostics due to language barriers, understanding test instructions, and results interpretation (32).

Our study identified several barriers to POC testing for STI/HIV testing at CP. One of the prominent challenges that emerged was the lack of privacy, which hindered open communication between clients and healthcare providers and may have negatively impacted testing uptake, consistent with previous studies (6). Participants proposed solutions such as setting up tents or designated safe spaces to ensure confidentiality and privacy, to foster open communication. Another significant barrier was the absence of adequate sanitary facilities, particularly affecting women who needed to collect samples. Many had to pay for nearby facilities, creating an inconvenience that discouraged testing. Another barrier identified was related to difficulties with self-sample collection, particularly with invasive methods, worsened by privacy concerns. Less invasive sample options, such as urine or saliva-based tests, were recommended to improve comfort and encourage testing, findings consistent with prior research (33, 34). Psychological barriers also played a significant role in limiting POC testing in our study. The fear of receiving a positive HIV result, often associated with stigma and shame, deterred people from POC testing in CPs. These findings are consistent with previous research, which shows that promoting pharmacy-based psychosocial support increased clients’ willingness to test and reduced anxiety (35), as well as enhanced engagement with follow-up care (36).

Concerns were also raised about pharmacists’ training and potential conflicts between professional ethics and commercial interests. Some participants questioned pharmacists’ competence in independent diagnostic sample collection and testing, which diminished confidence in the accuracy of POC testing in this study. In Uganda, diagnostic testing is currently restricted to authorized laboratory personnel under national regulatory frameworks. Limiting pharmacists’ involvement in sample collection and analysis (37). Participants suggested revising guidelines to allow specialized training and certification in sample collection and management. Others argued that testing should remain the responsibility of clinicians and laboratory professionals to ensure diagnostic integrity. Similar regulatory debates have been reported in South Africa, where pilot programs demonstrated that structured training, certification, and quality assurance mechanisms could enable safe expansion of pharmacy-based HIV testing while maintaining diagnostic integrity (29).

Lastly, concerns were raised about the limited scope and variable performance of current POC tests. Many healthcare providers and policy makers observed that, in practice, rapid tests are widely available for HIV, whereas POCTs for other non-HIV STIs remain largely unavailable. These concerns align with global evidence indicating that HIV rapid diagnostic tests are widely available and supported by clear regulatory and implementation frameworks, whereas reliable, affordable POCTs for other STIs remain limited in availability and demonstrate variable sensitivity and specificity in field settings (4). This diagnostic imbalance constrains the feasibility of integrating HIV/STI POCT delivery within CPs.

Our study had several strengths. The use of IDIs alongside an ongoing quantitative study of POC testing allowed for a nuanced exploration of facilitated participants’ real-time personal experiences and challenges in using POCTs. By involving pharmacy clients, healthcare providers, and policymakers, this study helped capture perspectives across multiple levels of HIV and STI management within communities, thereby enriching the depth and diversity of insights. Interviews were conducted in confidential settings by staff experienced in qualitative research and fluent in both English and Luganda, which helped minimize social desirability bias and ensured cultural and linguistic appropriateness.

However, the study had some limitations. The greatest limitation is that treatment and testing were offered for free, contrary to the actual practice. This shows that a co-funded approach with community pharmacies offering this service at no cost to clients would be acceptable, but it is unclear if clients themselves would pay more for a test and treatment as compared to empiric treatment. Data were collected only from urban and peri-urban pharmacies, which may limit how well the findings apply to rural settings where access, infrastructure, and awareness can be quite different. Because HIV and STI discussions are sensitive, social desirability bias might have affected some participant responses. The reported high acceptability of POCTs in community pharmacies could also have been influenced by the extra services offered during the study, such as free treatment, counselling, and partner notification, which are not usually available in regular pharmacy practice. Additionally, limited resources prevented back translation of interview transcripts, but all transcripts were reviewed for accuracy by trained research assistants to ensure the data’s validity and reliability.

### Recommendations

The analysis of this data highlighted many recommendations for integrating POC within CP. Firstly, there is some work needed to provide contextually appropriate support for this service. To address the lack of awareness of POC testing, comprehensive community sensitization campaigns will be needed using local media, flyers, and community outreaches to raise awareness of POC testing and its benefits. We found that low literacy levels among clients led to difficulties understanding instructions, and so we recommend developing user-friendly instructions in multiple languages and simplifying test instructions through visual aids, which is consistent with previous research We recommend integrating pharmacy-based psychosocial support into POC testing services after training pharmacists in counselling, stigma-reduction communication, test procedure, and results interpretation and linkage-to-care navigation to reduce anxiety and encourage adherence to POC testing. Regular capacity-building initiatives for CPs will strengthen adherence to community-level STI treatment guidelines, antimicrobial stewardship principles, and engagement in community outreach to raise awareness about STI/HIV testing.

At a policy level, we recommend that future policies consider integrating CPs into national SRH policies and clear ethical guidelines aligned with national HIV/STI management protocols, consistent with prior research (12, 38). For widespread rollout of CP STI testing, investment will be required to equip CP with basic sanitary infrastructure and private consultation areas to support private and safe self-sample collection, consistent with previous studies (29, 39). We recommend more research into the cost-effectiveness of STI treatment in CPs to inform national guidelines.

Research and development are needed for the design of POCTs that are less invasive, more user-friendly, and complemented by digital tools to aid in result interpretation, as documented by prior studies (40). Integrating innovative digital surveillance technologies, such as electronic medical records, mobile health applications, AI-powered analytics, and cloud-based data systems, can enhance antibiotic monitoring and real-time data collection, an area for future research.

## Conclusions

This study found that POC HIV/STI testing in CPs was perceived as highly acceptable and satisfactory among both pharmacy clients, healthcare workers, and policy makers, despite significant challenges related to awareness, privacy concerns, and inadequate infrastructure. Addressing these barriers through targeted policies, professional training, community education, and strengthened regulatory oversight could enhance the delivery and uptake of pharmacy-based testing services. Given their accessibility and trusted role in primary health care, CPs present a promising platform for expanding access to HIV and STI diagnostic services outside traditional clinical settings. Integrating these services into national health frameworks could not only improve early detection and treatment but also contribute valuable surveillance data to inform guideline development and strengthen public health outcomes.

## Data Availability

The IDI and KII guides are attached in the additional files. The source data is in recorded voice, which could break the anonymity of participants. The IDIs and KIIs were anonymized and transcribed. The data analyzed for this manuscript will not be made available online because of its sensitive nature, but the corresponding author can be contacted on a case-by-case basis to provide data following a reasonable request to

## ACKNOWLEDGEMENTS

The authors express their gratitude to all the study participants and the participating pharmacies in Kampala and Wakiso districts that allowed the study to be conducted within their premises. We are grateful to all the research assistants for their contribution in collecting data.

We acknowledge Dr. Caitlyn Kennedy from Johns Hopkins University for reviewing the manuscript draft and offering valuable feedback that contributed to its final version.

## AUTHORS’ CONTRIBUTIONS

All authors contributed to the study. AAO: Project administration, data collection, data analysis, draft manuscript writing. AAO is the guarantor; AK: conceptualized and designed the study, project supervision, manuscript reviews; RPR; Project supervision, study design, manuscript reviews; YCM: Funding acquisition, project supervision, manuscript reviews; AT: data analysis, manuscript reviews; MMH: Study design, project supervision, manuscript reviews; PK: Project supervision, manuscript reviews; JHM: Project support, manuscript reviews. All authors read and approved the final manuscript.

## ETHICS APPROVAL

The study was initially reviewed and approved by the Scientific Research Committee of the IDI. It was then approved by the Institutional Review Board (IRB) at The Joint Clinical Research Center (JC1820) in Uganda and the Johns Hopkins University (IRB00279241). The Uganda National Council of Science and Technology (HS1274ES) provided overall regulatory and administrative oversight and approval regarding the conduct of study activities.

## AVAILABILITY OF DATA AND MATERIALS

The IDI and KII guides are attached in the additional files. The source data is in recorded voice, which could break the anonymity of participants. The IDIs and KIIs were anonymized and transcribed. The data analyzed for this manuscript will not be made available online because of its sensitive nature, but the corresponding author can be contacted on a case-by-case basis to provide data following a reasonable request.

## ABBREVIATIONS

POCTs: Point of care Tests
IDI: In-depth Interviews
KII: Key informant interviews
CPs: Community pharmacies
UNCST: Uganda National Council of Science and Technology
REC: Research Ethics Committee
JCRC: Joint Clinical Research Center
IRB: Institutional Review Board
AMR: Antimicrobial Resistance
STI: Sexually Transmitted Infections
HIV: Human immunodeficiency Virus
MOH: Ministry of Health
RA: Research assistant

## REFERENCES

1. WHO. Global progress report on HIV, viral hepatitis and sexually transmitted infections, 2021: accountability for the global health sector strategies 2016–2021: actions for impact: web annex 2: data methods. 2021.

2. Eisinger RW, Erbelding E, Fauci AS. Refocusing Research on Sexually Transmitted Infections. The Journal of Infectious Diseases. 2020;222(9):1432–4.

3. Cohen MS, Council OD, Chen JS. Sexually transmitted infections and HIV in the era of antiretroviral treatment and prevention: the biologic basis for epidemiologic synergy. Journal of the International AIDS Society. 2019;22 Suppl 6(Suppl Suppl 6):e25355.

4. Wi TE, Ndowa FJ, Ferreyra C, Kelly-Cirino C, Taylor MM, Toskin I, et al. Diagnosing sexually transmitted infections in resource-constrained settings: challenges and ways forward. Journal of the International AIDS Society. 2019;22 Suppl 6(Suppl Suppl 6):e25343.

5. Igor T, Rosanna WP, David M, King H, Ronald B, James K, et al. Point-of-care tests for STIs: the way forward. Sexually Transmitted Infections. 2017;93(S4):S1.

6. Gubbins PO, Klepser ME, Dering-Anderson AM, Bauer KA, Darin KM, Klepser S, et al. Point-of-care testing for infectious diseases: Opportunities, barriers, and considerations in community pharmacy. Journal of the American Pharmacists Association. 2014;54(2):163–71.

7. Rompalo AM, Hsieh YH, Hogan T, Barnes M, Jett-Goheen M, Huppert JS, et al. Point-of-care tests for sexually transmissible infections: what do ‘end users’ want? Sexual health. 2013;10(6):541–5.

8. Hsieh YH, Hogan MT, Barnes M, Jett-Goheen M, Huppert J, Rompalo AM, et al. Perceptions of an ideal point-of-care test for sexually transmitted infections--a qualitative study of focus group discussions with medical providers. PloS one. 2010;5(11):e14144.

9. Morikawa E, Mudau M, Olivier D, de Vos L, Joseph Davey D, Price C, et al. Acceptability and Feasibility of Integrating Point-of-Care Diagnostic Testing of Sexually Transmitted Infections into a South African Antenatal Care Program for HIV-Infected Pregnant Women. Infectious Diseases in Obstetrics and Gynecology. 2018;2018:3946862.

10. Nuwaha F. Determinants of choosing public or private health care among patients with sexually transmitted infections in Uganda. Sexually transmitted diseases. 2006;33(7):422–7.

11. Ndayishimye S, Oladokun A, Mukanyangezi MF. User experiences of selfcare interventions for sexual and reproductive health services in community pharmacies in Rwanda: A qualitative study. Glob Public Health. 2024;19(1):2397691.

12. Navarrete J, Yuksel N, Schindel TJ, Hughes CA. Sexual and reproductive health services provided by community pharmacists: a scoping review. BMJ open. 2021;11(7):e047034.

13. Licensed Outlets Statistics [Internet]. 2025 [cited 24 February 2026]. Available from: https://www.nda.or.ug/licensed-outlets-statistics/.

14. Kamba PF, Mulangwa J, Kaggwa B, Kitutu FE, Sewankambo NK, Katabira ET, et al. Compliance of private pharmacies in Uganda with controlled prescription drugs regulations: a mixed-methods study. Substance Abuse Treatment, Prevention, and Policy. 2020;15(1):16.

15. Kiragga AN, Najjemba L, Galiwango R, Banturaki G, Munyiwra G, Iwumbwe I, et al. Community purchases of antimicrobials during the COVID-19 pandemic in Uganda: An increased risk for antimicrobial resistance. 2023;3(2):e0001579.

16. Sakeena MHF, Bennett AA, McLachlan AJ. Non-prescription sales of antimicrobial agents at community pharmacies in developing countries: a systematic review. International Journal of Antimicrobial Agents. 2018;52(6):771–82.

17. Kuo AP, Roche SD, Mugambi ML, Pintye J, Baeten JM, Bukusi E, et al. The effectiveness, feasibility and acceptability of HIV service delivery at private pharmacies in sub-Saharan Africa: a scoping review. J Int AIDS Soc. 2022;25(10):e26027.

18. Kyomugisha EL, Balaba M, Nakibuuka E, King R, Parkes-Ratanshi R. Stakeholder engagement to drive iterative software development of the ARTAccess web-based application for community pharmacy dispensing of anti-retroviral therapy. Medical Research Archives. 2023;11(1).

19. Meyerson BE, Ryder PT, Richey-Smith C. Achieving pharmacy-based public health: a call for public health engagement. Public health reports (Washington, DC: 1974). 2013;128(3):140–3.

20. Rosenthal M, Austin Z, Tsuyuki RT. Are Pharmacists the Ultimate Barrier to Pharmacy Practice Change? 2010;143(1):37–42.

21. Wiedenmayer K, Summers RS, Mackie CA, Gous AG, Everard M, Tromp D, et al. Developing pharmacy practice: a focus on patient care: handbook. World Health Organization; 2006.

22. Kiragga AN, Onzia A, Nakate V, Bagaya I, Natuha E, Mande E, et al. Community pharmacies: Key players in point-of-care diagnostics for STI screening in Africa. 2024;19(12):e0315191.

23. Bwanika Naggirinya A, Meya DB, Nabaggala MS, Banturaki G, Kiragga A, Rujumba J, et al. Effectiveness of interactive voice response-call for life mHealth tool on adherence to anti-retroviral therapy among young people living with HIV: A randomized trial in Uganda. 2024;19(11):e0308923.

24. Uganda Bureau of Statistics. Statistical abstract. UBOS; 2020 2020.

25. Sekhon M, Cartwright M, Francis JJJBhsr. Acceptability of healthcare interventions: an overview of reviews and development of a theoretical framework. 2017;17:1–13.

26. Ministry of Health. Uganda Clinical Guidelines 2023. 2023.

27. Younas A, Fàbregues S, Durante A, Escalante EL, Inayat S, Ali P. Proposing the “MIRACLE” narrative framework for providing thick description in qualitative research. International Journal of Qualitative Methods. 2023;22:16094069221147162.

28. Tong A, Sainsbury P, Craig J. Consolidated criteria for reporting qualitative research (COREQ): a 32-item checklist for interviews and focus groups. Int J Qual Health Care. 2007;19(6):349–57.

29. Nyamuzihwa T, Tembo A, Martyn N, Venter F, Maimin J, Houghton J, et al. The South African community pharmacy sector—an untapped reservoir for delivering HIV services. 2023;Volume 5 - 2023.

30. Martin K, Wenlock R, Roper T, Butler C, Vera JH. Facilitators and barriers to point-of-care testing for sexually transmitted infections in low- and middle-income countries: a scoping review. BMC Infectious Diseases. 2022;22(1):561.

31. Nnko S, Changalucha J, Mosha J, Bunga C, Wamoyi J, Peeling R, et al. Perceptions, attitude and uptake of rapid syphilis testing services in antenatal clinics in North-Western Tanzania. 2016;31(5):667–73.

32. Engel N, Ganesh G, Patil M, Yellappa V, Vadnais C, Pai NP, et al. Point-of-care testing in India: missed opportunities to realize the true potential of point-of-care testing programs. BMC Health Services Research. 2015;15(1):550.

33. Ogale YP, Grabowski MK, Nabakka P, Ddaaki W, Nakubulwa R, Nakyanjo N, et al. Self-collected samples as an additional option for STI testing in low-resource settings: a qualitative study of acceptability among adults in Rakai, Uganda. BMJ open. 2023;13(11):e073241.

34. Melendez JH, Muñiz Tirado A, Onzia A, Mande E, Hardick JP, Parkes-Ratanshi R, et al. Self-collected penile-meatal swabs are suitable for the detection of STIs in Ugandan men with high rates of STI coinfections. Sex Transm Infect. 2025;101(4):247–51.

35. Rietmeijer CA. Risk reduction counselling for prevention of sexually transmitted infections: how it works and how to make it work. Sex Transm Infect. 2007;83(1):2–9.

36. Lalla-Edward ST, Venter WDF. Feasibility and impact of community pharmacy and novel pick-up points for antiretroviral therapy pre-exposure prophylaxis initiation and continuation in low and middle-income countries. Current HIV/AIDS Reports. 2025;22(1):2.

37. Uganda PSO. Code of Conduct for Pharmacists in Uganda. In: Health, editor. Kampala Uganda 2014. p. 31.

38. Raju R, Srinivas SC, Siddalingegowda SM, Vaidya R, Gharat M, Kumar TMP. Community pharmacists as antimicrobial resistance stewards: a narrative review on their contributions and challenges in low- and middle-income countries. Journal of pharmacy & pharmaceutical sciences: a publication of the Canadian Society for Pharmaceutical Sciences, Societe canadienne des sciences pharmaceutiques. 2024;27:12721.

39. Bernknopf AC, Bronz HE, Klepser MEJJotACoCP. Point-of-care testing in Community pharmacies to expand access to treatment for sexually transmitted infections. 2025.

40. Wosny M, Strasser LM, Hastings J. Experience of Health Care Professionals Using Digital Tools in the Hospital: Qualitative Systematic Review. JMIR human factors. 2023;10:e50357.

